# Diffusion Weighted MRI could precisely predict the pTERT mutation status of GBM using a residual convolutional neural network

**DOI:** 10.1101/2022.12.28.22283931

**Authors:** Congman Hu, Ke Fang, Quan Du, Jiarui Chen, Lin Wang, Lanjuan Li, Jianmin Zhang, Ruiliang Bai, Yongjie Wang

## Abstract

**Background:** Telomerase reverse transcriptase promoter (pTERT) mutation status plays a key role in the decision-making and prognosis prediction of glioblastoma (GBM). The purpose of this study was to assess the prediction value of diffusion-weighted imaging (DWI) in the pTERT mutation status of GBM

**Methods:** MR imaging data and molecular information of 266 patients with GBM were obtained from the Second Affiliated Hospital of Zhejiang University (n=266). We trained the same residual convolutional neural network (ResNet) for each MR modality, including structural MRIs (T1-weighted, T2-weighted, contrast enhanced T1-weighted) and DWI and its associated ADC map, and their combinations to compare the predictive capacities between DWI and conventional structural MRI. Moreover, we explored the effect of different Region of interests (ROIs) on the outcome of pTERT mutation status prediction: entire tumor (solid tumor, edema and cystic regions), tumor core (solid tumor), and enhanced tumor.

**Results:** Structural MRI modalities and their combination performed poorly in predicting the pTERT mutation status (accuracy, 51-54%, AUC, 0.545-0.571), while DWI in combination with its ADC maps yielded the best predictive performance (accuracy = 85.2%, AUC= 0.934). The further including of radiological and clinical characteristics could not further improve the predictive performance of pTERT mutation status. Among the three ROI selections, the entire tumor volume yielded the best prediction performance.

**Conclusion:** DWI and its associated ADC maps shows promising prediction value in the pTERT mutation in GBM and are suggested to be included into the MRI protocol of GBM in clinical practice.

**Key Points:**

- The ResNet model constructed by radiomics provided great help for the prediction of pTERT mutation in glioblastoma.
- In the ResNet prediction model, conventional structural MRI was of little value while DWI and its associated ADC maps shows excellent value.
- The model using the whole tumor as ROI showed best predictive capacity and potentiality for future clinical application.

## Introduction

High-grade glioma is the most common primary brain tumor in adults and accounts for 81% of brain malignancies. As the most frequent and lethal subtype, glioblastoma (GBM) has an extremely poor prognosis, with a median survival of around 15 months after radical resection followed by concurrent chemoradiotherapy and adjuvant chemotherapy with temozolomide (TMZ)[1]. Studies had proved several genetic markers as important indicators for treatment response and overall survival, including isocitrate dehydrogenase (IDH)1/2 mutation, 1p/19q co-deletion, mutations in the telomerase reverse transcriptase promoter (pTERT), methylation of O6-methylguanine-DNA methyltransferase (MGMT) promotor methylation status, et al.[2][3]. As a result, molecular pathology has been incorporated into the new integrated diagnosis of WHO[4].

Mutations in pTERT result in increased expression of telomeres, essential for cancer cells to avoid aging and maintain proliferative potential, and serve as an important biomarker in glioma diagnosis[5]. Studies have shown that pTERT was mutated in almost 80% of primary GBM, which was more common than IDH 1/2 mutations[6]. Additionally, pTERT mutations have shown to be a distinct, independent, and superior prognostic marker in adult GBMs[2][7][8]. pTERT mutations in GBM were associated with poorer prognosis, and could significantly benefit from more aggressive surgical and chemotherapy strategies[2]. As a result, considerable efforts have been made to develop anti-cancer drugs targeting telomerase, and many TERT peptide vaccines are undergoing clinical trials[5][9][10]. Above all, the pTERT mutations status is a key biomarker in GBM as well as future therapeutic targets[11]. Preoperative prediction of pTERT mutations can help with more accurate treatment decision making.

At present, pathological diagnosis regarding underlying genetic and molecular alterations of gliomas is still based on post-operational molecular analysis of tumor tissue obtained by means of surgical resection or biopsy, which carries drawbacks of sampling error, invasive nature, money and time-consuming[12][13]. MRI radiomics coupled with either machine learning or deep learning provide a noninvasive way to assess both global and regional characterization of GBM[14], and have been successfully applied for the accurate prediction of key molecular markers such as IDH[15][16][17][18], 1p/19q[19] and MGMT[20] promoter methylation[20]. However, non-invasive prediction of pTERT mutation status in GBMs using MRI data still remains a challenge. Tunc F Ersoy et al. reported that conventional structural MRI biomarkers (i.e., T1-weighted image (T1WI), T2-weighted image (T2WI), and contrast enhanced T1WI (CE-T1)) lacks the capability to forecast pTERT mutation[21]. Similarly, Jana Ivanidze et al. concluded that pTERT mutation lacked typical imaging features in conventional structural MRI [22]. Tian et al. used radiomics analysis based on multi-modality structural MRI, and also failed to establish a predictive model for pTERT mutation status in GBM[23]. Overall, the conventional structural MRI lacks the capability to predict pTERT mutation status in GBM.

Diffusion weighted image (DWI) and its associated apparent diffusive coefficient (ADC) value is a well-established functional MRI method in clinic and shows potentials in characterizing tissue microstructure in tumor [24]. Successful applications of DWI in the predicting of glioma grading[24][25][26][27][28], diagnosis [29], and prognostic value[30]. But, whether DWI modality has prediction value of pTERT mutation is still largely unknown. As DWI could be easily implemented in clinical practice in adjunction to conventional structural MRI, it is highly desired to explore the prediction value of DWI in pTERT mutation.

In this study, we aim to study the capacity of DWI in the prediction of pTERT mutation in GBM. In predicting the genetic and molecular biology of tumors based on MRI, convolutional neural networks (CNNs) show effective performance and outperform traditional machine learning methods[31]. Among the various CNNs, residual convolutional neural network (ResNet) exploits extensive stacks of learnable convolutional filters and had been shown to be one of the most efficacious for medical imaging[32]. It had a good effect on the prediction of IDH status, MGMT methylation status and other gene mutations of glioma[20][16]. Here, we tried to explore the prediction of pTERT mutation status in GBMs using ResNet, which was trained on randomly selected dataset from each MRI modalities to compare their prediction value, namely the conventional structural MRI (T1-weighted image (T1WI), T2-weighted image (T2WI), and contrast enhanced T1WI (CE-T1)) and DWI techniques (DWI with its associated ADC maps). In addition, the prediction value of radiological characteristics including location, edema and necrosis area, and patient’s age were also explored. At last, the influence of region of interest (ROI) selection on the predictive efficacy was also examined.

## Materials and Methods

### Patient selection

We retrospectively searched the electronic database of GBM patients operated in the Neurosurgery Department of the Second Affiliated Hospital of Zhejiang University (2^nd^ HZJU) between March 2018 and October 2021 (n=386) (**Figure** 1). MRI, primary patient demographic data (age, gender, et al.), pathological information including hemotoxin-eosin staining, immunohistochemistry analysis and molecular genotyping data were obtained and reviewed. The inclusion criteria were as follows: 1) pathologically confirmed primary GBM according to 2016 WHO diagnostic criteria; 2) pTERT mutations including C228T and C250T confirmed by Quantitative Real-time PCR(qPCR)[33], sanger sequencing following standard PCR amplification[34], or whole exon sequencing based on next-generation sequencing (NGS)[35]; 3) MRI within 1 month before operation, including T1WI, T2WI, CE-T1, with or without DWI; 4) age≥18 years. The exclusion criteria were as follows: 1) significant artifacts resulting in low imaging quality; 2) previous history of biopsy, surgery, radiotherapy or chemotherapy for brain lesions of any kind; 3) coexistence of other intracranial lesions. Our final patient cohort included 266 patients, with DWI information available for 147 patients. The study was approved by the institutional review board of 2^nd^ HZJU. The requirement for informed consents was waived.

**Figure 1.**
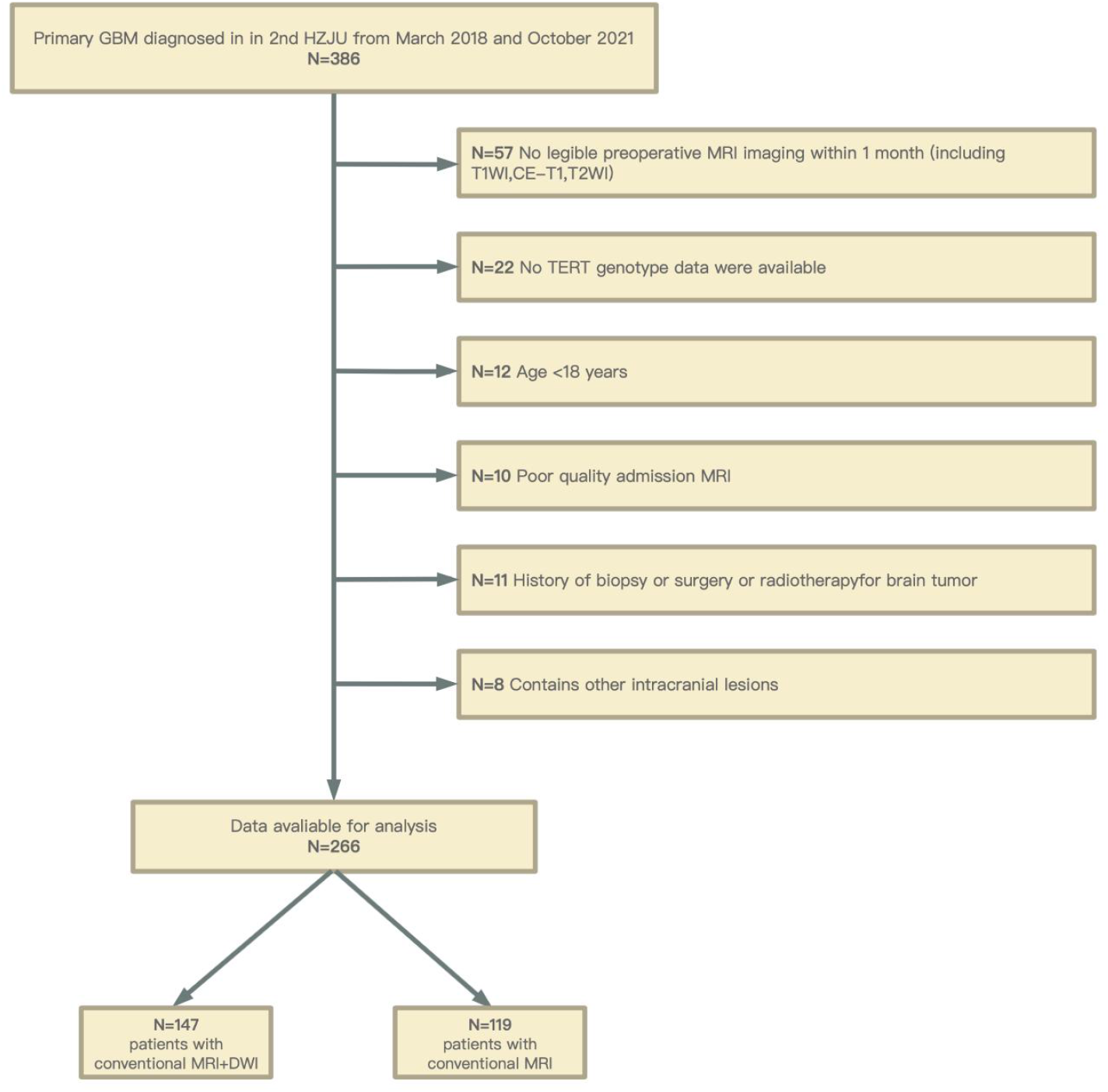
Flow diagram of the study population. 2^nd^ HJZU, the Second Affiliated Hospital of Zhejiang University; TERT, Telomerase reverse transcriptase.

### Study Design

The study design was summarized in **Figure** 2. MR Imaging protocols included T1WI, T2WI, CE-T1, with or without DWI data. They were registered using T1WI as the standard. The image was normalized to signal intensity and resampled to a size of 448×448 voxels (**Figure** 2.A). The input tumor region was manually delineated as different ROIs, and finally the ResNet-based binary classifier was trained to predict the pTERT mutation status (**Figure** 2.C). In order to explore the predictive value of DWI, we trained ResNet models based on different MRI modality, and group

**Figure 2.**
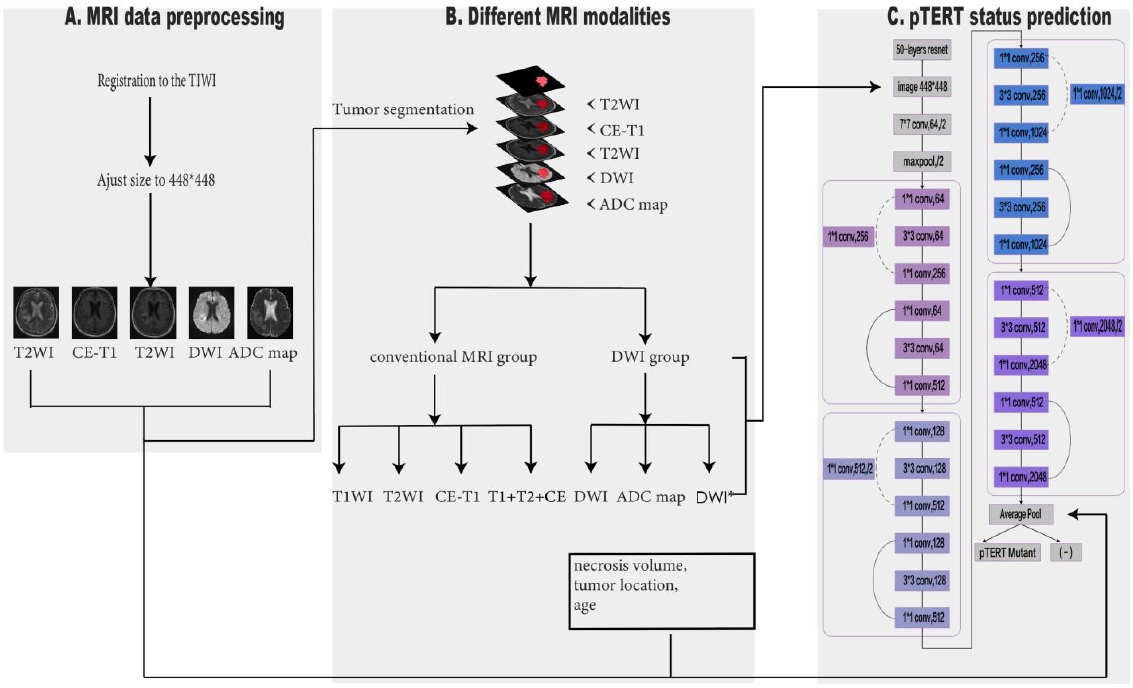
Flow diagram of pTERT mutation status prediction. **A**, Image preprocessing steps in our proposed approach. **B**, Comparison of different MRI modalities. **C**, A modified 50-layer residual neural network architecture was used to predict pTERT mutation status. Conv and pool stand for convolutional and pooling. The pooling size used was 2 (denoted by “/2.”). The first box (“7 × 7 conv, 64”) meant that the convolutional kernel size was 7 × 7 with 64 filters. We then explicitly described the following layer as a 2 × 2 pooling layer, but elsewhere in this figure we used the shorthand of “/2” in the box showing the layer. Solid lines (—) indicate identity and dashed lines (- - -) indicate cross-residual weighted connections. DWI*, DWI and its associated ADC map.

### Imaging acquisition and data processing

comparison was performed (**Figure** 2.B). Age and other clinical characteristics were added to the optimization model. All the patients underwent conventional structural MR and/or DWI (with b values of 0 and 1000 s/mm2). MRI was performed on a 1.5T scanner. The detailed imaging parameters are as following: T2WI with TR = 3500-4000 ms, TE = 96-107 ms, section thickness of 6mm, in-plane resolution 4mm×4mm; T1WI with TR = 1800 ms, TE = 7.5 ms, section thickness = 6mm, in-plane resolution 4mm×4mm; CE-T1 images were obtained after injection of 0.1 mmol/kg gadolinium with the same imaging parameter as T1WI; DWI images were obtained prior to injection of the contrast agent, with TR = 10,500 ms, TE = 96 ms, section thickness = 6mm, in-plane resolution 1.5mm×1.5mm.

ADC map was calculated from the DWI results, using FSL [FMRIB Software Library v5.0.2.2] with the following equation: 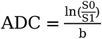, where S1 was the acquired signal (averaged over 3 directions) under b = 1000 s/mm^2^, and S0 was the signal acquired without diffusion gradients. In each subject, the other MRI modalities were rigidly registered to the T1WI and the image intensity of MRI (except for ADC maps) were normalized before further processing and statistics.

### Region of interests (ROIs) selection

Two neuroradiologists manually marked the entire tumor (tumor with edema area), the tumor core, the enhanced portion and the necrotic portion (cystic component), which were labeled as entire tumor, 2, 3 and 4 respectively (**Figure** 3). entire tumor was selected as abnormal area in T2WI. tumor core was drawn by excluding edema signal on T2WI. enhanced tumor was selected based on CE-T1. We used the strict criteria of “clear” necrosis as the non-enhanced portion on CE-T1 images, with similar intensity as CSF on T2WI. Itk-snap (version 3.8), a user-driven manual active contour segmentation tool, was used to segment tumor volumes. After secondary correction by a neuroradiologist, the segmented ROI was overlapped on T1WI, T2WI, CE-T1, DWI and ADC map. These ROIs were used to generate piece-by-piece cropped images of tumors in all modes. To facilitate network training, random number replacement was used to erase the image information outside the ROI so that each image was subsequently adjusted to 448×448 voxels input.

**Figure 3.**
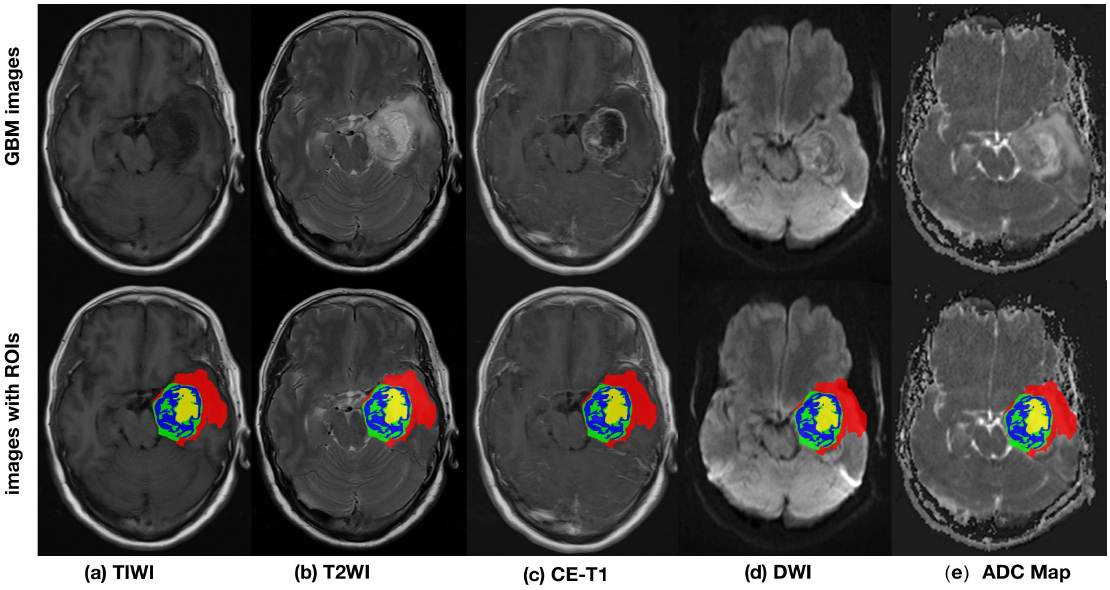
Segmentation result from a training image of GBM, overlapping on T1WI(a), T2WI(b), CE-T1(c), DWI(d) and ADC map(e). Red: edema; Green: non-enhancing tumor solid portion. Blue: enhanced portion; Yellow: cystic or necrotic portion. **entire tumor**: red, green, blue, and yellow. **tumor core**: green, blue. **enhanced tumor**: blue.

In order to enhance the model robustness, we performed data enhancement steps including horizontal inversion, vertical inversion and random translational rotation of the ROI. In addition, normal ROIs were generated from 97 patients without midline involvement by mirroring of the tumor ROIs onto the contralateral side, and were also incorporated into the training model. Enhancements were performed only on the training set, not on the test set.

### Network scheme

In this study, a ResNet structure was selected for pTERT mutation status prediction using different MR modalities. The input image was transformed through a series of chained convolutional layers that resulted in an output vector of class probabilities. As shown in **Figure** 2.C, our ResNet model was derived from a 50-layer residual network architecture and divided into five stages. The structure of stage 0 was relatively simple and could be regarded as input preprocessing. The last four stages were composed of relatively similar bottleneck structures. The features extracted from the remaining blocks were pooled using average pooling and sent to the output layer, which used the sigmoid activation function for prediction.

### Model training

The images input to train the ResNet model were comprised of axial T1WI, T2WI, CE-T1, DWI and associated ADC map with tumor masks of 448 × 448 voxels. The predictive capacity of different MRI modalities namely DWI group (DWI, ADC map, DWI+ADC map (DWI*)) and conventional MRI group (T1WI, T2WI, CE-T1, T1WI+T2WI+CE-T1), on pTERT mutation status were compared. A total of 266 patients were entered into ResNet. The total number of patient samples were 2081 for DWI group and 3116 for conventional MRI group. For DWI, 30 randomly chosen patients who contributed 262 samples per sequence, that were never seen by the model during training were used as a test set. The rest of patients was input to the network as a training set. For conventional structural MRI, 60 patients who contributed 660 samples per sequence were randomly selected as a test group.

Probabilistic analysis was used to determine the pTERT mutation status of tumor slice. For a specific tumor slice in the test set, the probabilities of TERT-W, TERT-M and normal were generated, with sum of 1. We compared the probabilities of TERT-W and TERT-M and chose the higher one to denote pTERT mutation status of each tumor slice. The pTERT mutation status of each patient in the test group was determined based on whether the proportion of positive pTERT mutated slices were more than 50%. The sizes of experimental data were summarized in Table 1.

**Table 1.** ResNet registration dataset

|  |  | Training set |  | Test set |  |
| --- | --- | --- | --- | --- | --- |
|  |  | Patients | Slices | Patients | Slices |
| DWI group | normal | 97 | 740 | 0 | 0 |
|  | TERT-M | 67 | 631 | 15 | 139 |
|  | TERT-W | 50 | 448 | 15 | 123 |
|  | Total | 117 | 1079 | 30 | 262 |
| Conventional MRI group | normal | 97 | 740 | 0 | 0 |
|  | TERT-M | 114 | 1075 | 30 | 266 |
|  | TERT-W | 92 | 808 | 30 | 237 |
|  | total | 206 | 1883 | 60 | 503 |

We further compared the effects of different ROI selection on the prediction of tumor pTERT mutation status. The mask data of entire tumor, tumor core, and enhanced tumor were input to build the predictive models. The ResNet with DWI* was selected for further comparison across different ROIs.

### TERT-related radiological and clinical characteristics

Previous studies have shown that the pTERT mutation may be related to age[36], location[37], and necrosis volume[38]. Age, gender, primary tumor site, tumor laterality at diagnosis were extracted from the raw data. Area of tumor necrosis and edema, the percentage of necrosis and edema were extracted from the processed MRI data. Primary sites were classified as frontal lobe, parietal lobe, temporal lobe, occipital lobe, insular lobe, basal ganglia, brain stem and cerebellum. Tumor laterality was defined as a ternary variable, left of center line, right of center line, left and right of center line. The factors associated with pTERT mutation in GBM were analyzed using binary logic univariate regression, except laterality which were analyzed by multivariate logistic regression, and the variables with significant difference were input into a support Vector Mac (SVM) model[38]. the results of the ResNet and the factors included were input into the trained classification model based on SVM algorithm.

### Prediction performance evaluation

The performance of the model was evaluated by evaluating the predictive accuracy of the test sets. Moreover, receiver operating characteristics (ROC) analysis was performed using sigmoid probabilities to obtain ROC curves and calculate the area under curve (AUC). The optimal dataset selection and architecture of the model were determined based on the AUC obtained from the test dataset.

### Statistical Analysis

Data were presented as the number of patients with percentage for categorical variables, means ± standard deviation for parametric continuous variables. All statistical analyses were performed using SPSS Version 25 (IBM Corp, Armonk, New York, USA). A *p*-value of <0.05 was considered to indicate a statistically significant difference.

## Result

### Patient characteristics

A total of 14562 samples were generated from 266 GBM patients (168 males, 98 females), with age of 55.2 ± 14.8 years. 124 patients (46.6%) belonged to TERT wild-type (TERT-W) group (78 males, 48 females), with age of 53 ± 16 (SD) years. 142 patients (53.4%) belonged to TERT mutation-type (TERT-M) group (90 males, 52 females) with age of 57 ± 15 years. Basic information for all patients was summarized in Supplementary Materials 1.

As shown in Table 2, we used univariate analyses to screen the potential independent prognostic factors of pTERT. The variables that were not validated as independent prognostic factors included gender, laterality of the tumor, percentage of tumor edema, and percentage of tumor necrosis. Age at diagnosis (95% CI 1.007-1.043, *p*=0.049), necrosis volume (95% CI 1.366-7.929, *p*=0.042), occipital lobe (95% CI 1.366-7.929, *p*=0.008) and brain stem (95% CI 0.029=0.696, *p*=0.016) were the significant features of TERT-M GBM.

**Table 2.**
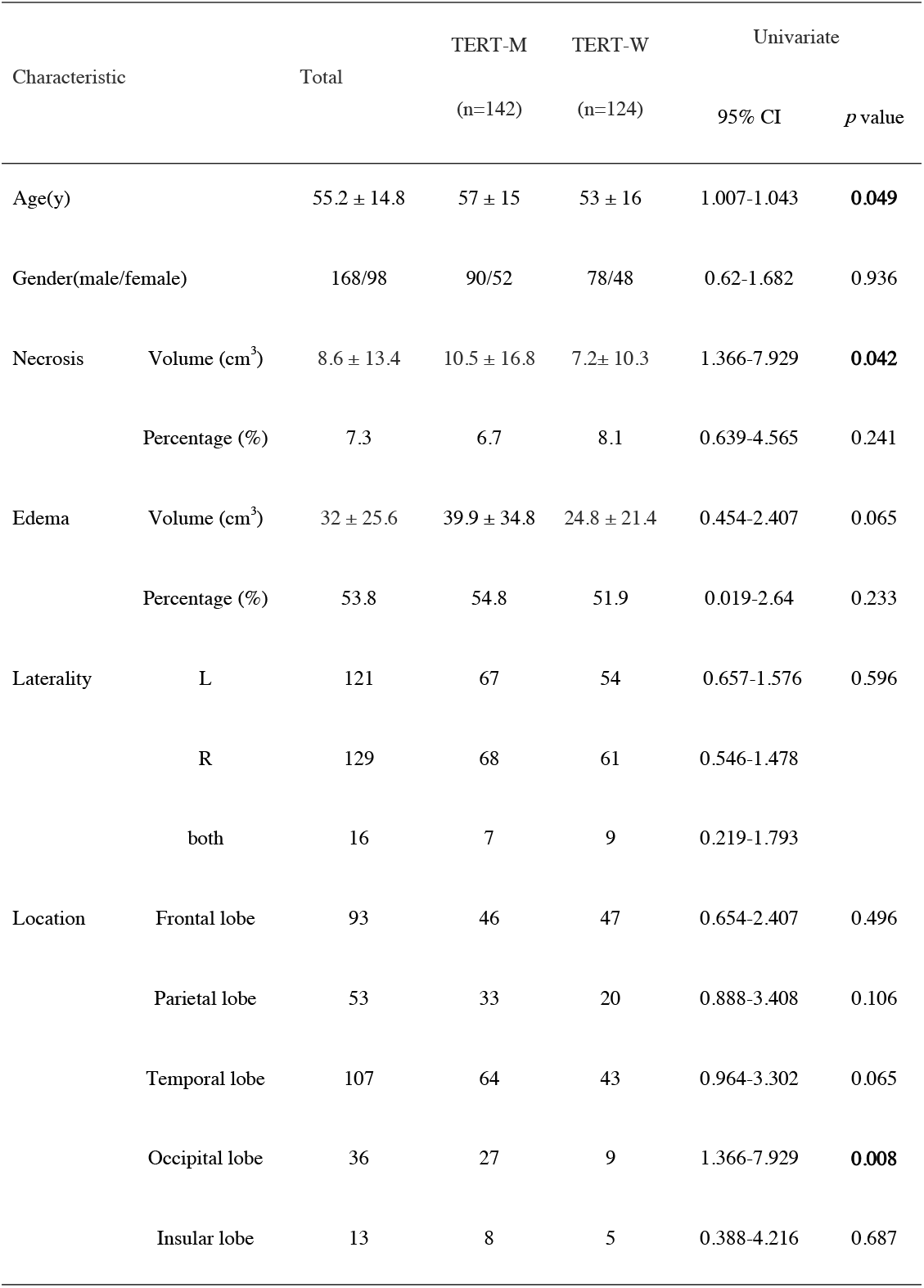
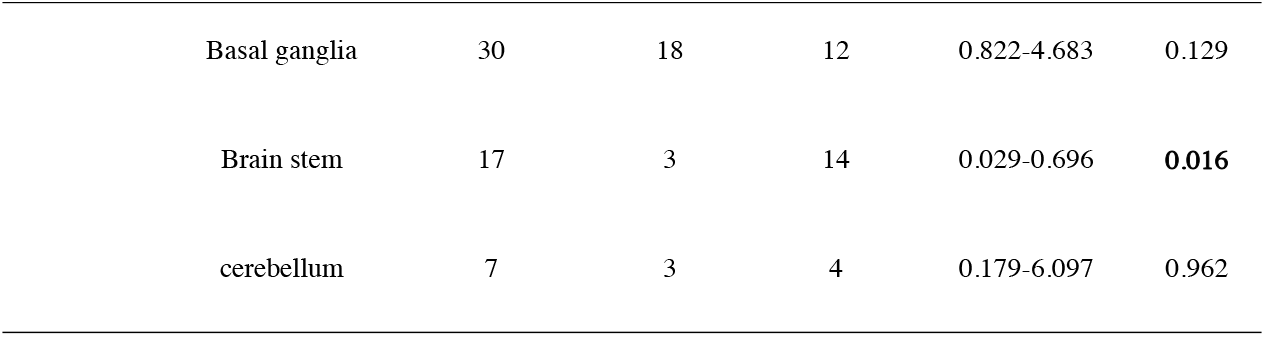
Univariate analysis of radiological and clinical characteristics in GBM with different TERT mutation status L, left; R, right

### Structural MRI vs DWI

Table 3 shows the prediction performance of the ResNet model trained from different MRI modalities with entire tumor. The prediction accuracies of T1WI, T2WI, CE-T1 and the combination of all structural MRI in the testing dataset were 50.38%, 51.91%, 51.53 and 50.10% (AUCs=0.547, 0.564, 0.571 and 0.527), respectively. In the DWI group, the prediction accuracy of DWI alone was only 54.58% (AUC =0.626), while the prediction accuracy of ResNet corresponding to ADC map reached 78.63% (AUC=0.865). The ResNet using DWI* (DWI and its associated ADC) data together showed the best performance in predicting pTERT status of each MRI slice with 85.20% accuracy (AUC=0.934, Table 3). The patient-based predictive accuracy was also excellent, being 86.6% (26/30) in the test group (The specific experimental results are shown in Supplementary Materials **2**). Overall, ResNet using DWI* data had a significant advantage over the conventional structural group in terms of predictive accuracy and AUC in testing set and showed best performance (**Figure** 4.A).

**Table 3.** Predictive performance of different MRI modalities of ResNet prediction model, SVM classifier model and a combination model DWI*, DWI+ADC; characteristics*: SVM clarifier ; SVM, support Vector Mac;

|  |  | Accuracy% | AUC |
| --- | --- | --- | --- |
| Per slice |  |  |  |
| DWI* group | DWI | 54.58 | 0.626 |
|  | ADC | 78.63 | 0.865 |
|  | DWI+ADC | **85.20** | **0.934** |
| Conventional MRI group | T1WI | 50.38 | 0.547 |
|  | T2WI | **51.91** | 0.564 |
|  | CE-T1 | 51.53 | **0.571** |
|  | T1+T2+CE-T1 | 50.10 | 0.545 |
| DWI*+Conventional structural MRI | T1+T2+CE-T1+DWI+ADC | 75.57 | 0.818 |
| characteristics* | necrosis volume,tumor location,age | 70.91 | 0.735 |
| DWI*+characteristics* | DWI+ADC+necrosis volume,tumor location,age | 79.92 | 0.898 |
| Per patient | DWI* | 86.67 |  |

**Figure 4.**
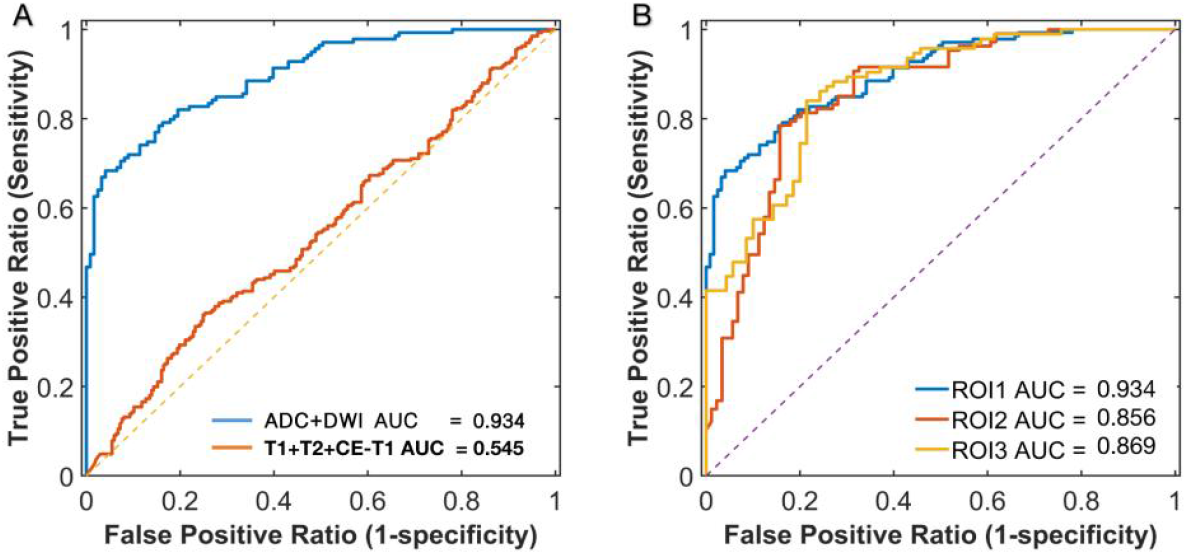
ROC plots of different ResNet models. A, the ROC plots comparing ResNet models using DWI* data and the combined conventional structural MRI (T1WI, T2WI and CE-T1) in the prediction of pTERT mutation status. B, the ROC plots of ResNet models using DWI* data for three different ROIs. ROC, receiver operating characteristic;

### Prediction performance of different ROIs

ResNet using DWI* data was subsequently tested using different ROIs. Table 4 and Figure 4.B summarized the accuracies and ROC analyses of different ROIs in distinguishing pTERT-mutated events from negative events. ResNet using the entire tumor performed best with an accuracy of 85.20% (AUC =0.934), while that using tumor core and enhanced tumor exhibited accuracies of 78.06% (AUC=0.856) and 81.70% (AUC=0.869) respectively.

**Table 4.** Predictive performance of different ROIs of ResNet prediction model using DWI* data ROI-DWI*, different ROIs of ResNet prediction model base on DWI* date

| ROI-DWI* | ROI | Accuracy% | AUC |
| --- | --- | --- | --- |
|  | entire tumor | 85.20 | 0.934 |
|  | tumor core | 78.06 | 0.856 |
|  | enhanced tumor | 81.70 | 0.869 |

### The prediction value of clinical characteristics

As indicated in Table 2, age, necrosis volume, and tumor location were significantly different between the TERT-M and the TERT-W groups (*p*<0.05) in the training cohort by univariate analysis. Here we further explored the prediction value of these clinical characteristics in pTERT mutation. Using SVM classifier with all the three clinical characteristics as inputs, the clinical characteristics had a relatively low prediction accuracy of 70.91% (AUC = 0.735). The further addition of these three clinical characteristics into DWI* data by building a model combining SVM classifier and RestNet (see methods in Supplementary Materials) didn’t make the predictive performance better than RestNet model based DWI* data, suggesting DWI* data itself has enough prediction value and the further addition of clinical characteristics is not necessary.

## Discussion

Pre-treatment identification of pTERT mutation status is important in clinical decision making because it can guide diagnosis, surgical strategy and chemotherapy selection for GBM. In this study, we demonstrated the value of DWI and its associated ADC map in predicting pTERT mutation status through deep learning and constructed a pTERT mutation status prediction model for GBMs. We further found that the ResNet model performed best using the entire tumor volume and the further inclusion of clinical characteristics into the prediction model was not necessary. Based on these results, it is highly suggested to include DWI into the clinical management of GBM.

In the ResNet models based on the entire tumor volume, the best prediction accuracy from the DWI group was 85.20% (AUC = 0.934), while the best prediction accuracy in the conventional MRI group was only 51.53% (AUC = 0.571). Previous studies had shown that it was difficult to find predictive radiological markers for pTERT mutation status using conventional structural, and our results also indicated that conventional structural MRI has limited value in pTERT mutation prediction[39][40]. However, few studies used diffusion-based MRI techniques to predict pTERT mutation status in glioma. Here, we found a significant improvement in the prediction performance of ResNet models using DWI and its associated ADC map (accuracy 78.63%, AUC 0.865). Because telomere maintenance is necessary to sustain infinite cell proliferation, pTERT mutations may lead to cell growth and proliferation, hence higher cell density, which is also associated with tumor aggressiveness[41][42][43]. A number of investigators showed an inverse linear relationship between ADC values and cell density in cerebral tumors [44][45]. Therefore, we speculated that the status of pTERT might be correlated with the ADC value of the tumor. However, Jana Ivanidze et al. found no significant difference between TERT-W and TERT-M tumors in histogram analysis of ADC values of GBM[40]. K. Yamashita et al. used diffusion-based MR images to predict pTERT mutations in patients with IDH wild-type GBM, similarly showing no difference in ADC values between TERT-W and TERT-M GBM[38]. This may be due to spatial and temporal heterogeneity based on tumor cell structure, vasogenic edema, degenerative changes (hemorrhage, cystic or mucinous degeneration), and/or compression of normal structures that destroys normal anatomical structures. As a result, signal changes may be additive or cancel each other out when evaluated using average ADC values[46]. The ResNet model, however, could completely analyze the distribution and texture changes of DWI signals and ADC values throughout tumor volumes and surrounding structures, and displayed significant advantages over the traditional ADC value analysis. In addition, we constructed a ResNet model combining DWI and ADC map, which resulted in further improvement of the prediction performance, with accuracy of 85.2% and AUC of 93.4%.

In this study, we found ResNet using the entire tumor achieved the best predictive performance. Interestingly ROI containing only enhanced tumor also showed acceptable results with predictive accuracy of 81.7% and AUC of 0.869 in the test set. The enhanced area is the core region of GBM that contains the most compact cellular and vascular architecture compared with the invasive margin. Moreover, the void signal in the center of the enhanced region commonly represents the necrotic area, which is proved to be correlated with pTERT mutation[38]. Therefore, using enhanced tumor as ROI might provide texture information most relevant with pTERT mutational status. Since it is easiest to distinguish enhanced tumor from other component, we can depict this ROI for preoperative screening. For tumor with tiny enhancement, we can change into ROI of the whole tumor for predictive analysis.

In our cohort, patient age, necrosis volume and tumor location are important features of TERT-M GBM but their prediction performance is worse than that of DWI* data. The SVM classifier model including these clinical features achieved a moderate predictive performance (accuracy=70.9%, AUC=0.735), which was similar with the results obtained by K Yamashita et al.[38]. However, the further incorporation of this SVM classifier model into our ResNet model using DWI* data doesn’t further improve the prediction performance of the latter, suggesting that it is not necessary to include these clinical characteristics in the prediction model using DWI* data.

Our study had several limitations. Firstly, this study is still a small-sample study. Due to the small sample size, most of our results was based on the cross-sectional slices rather than the whole tumor volume. Though the prediction performance was also tested on the patient-level using the 3D tumor volume and our results demonstrated the special value of DWI*, it is still highly desired to further test the results in a large sample using the 3D tumor volume in future. Moreover, our study was a retrospective study, and only focused on glioma diagnosed as WHO IV GBM by pathological analysis after surgery. In the future, we will try to explore the application of DWI technique in predicting pTERT mutation in glioma with lower WHO grade.

## Conclusion

In conclusion, we first proved the value of DWI techniques in predicting pTERT mutation status of GBM as compared with conventional structural MRI modality. Based on ResNet prediction model, DWI combined with its derived ADC map showed greatest value in the pTERT prediction of GBM. The model using the whole tumor as ROI showed best predictive capacity and potentiality for future clinical application.

## Data Availability

All data produced in the present study are available upon reasonable request to the authors

## Data availability

Data supporting the results of this study are available from the corresponding author (Ruiliang Bai and Yongjie Wang). The data is not public because it contains information that could compromise the privacy of study participants.

## Declarations

### Ethics approval and consent to participate

Approval of the study was obtained from the Ethics Committee of the Second Affiliated Hospital of Zhejiang University, School of Medicine.

### Informed consent

Informed written consent was obtained from the patients for publication of this cohort study and accompanying images.

## Acknowledgement

The authors have no acknowledgement to disclose.

## Funding

The whole work was supported by grants from the National Natural Science Foundation of China (81502139).

Clinical Research Center for Neurological Diseases of Zhejiang Province(2021E50006).

